# Hospital length of stay and discharge type prediction using deep learning

**DOI:** 10.1101/2023.07.24.23293092

**Authors:** Vikas Ramachandra, Prateek Sanghi

**Affiliations:** Onward Health

## Abstract

The length of hospital stay (LOS) and the type of discharge are important indicators of how well care is provided at a hospital. The purpose of this study is to leverage in-patient data collected at the hospital to help determine the factors that influence the length of hospital stay and type of discharge. Our research focuses on estimating if the person survived or not after they were admitted to the hospital, as well as the type of discharge. The study uses a retrospective design and examines information from hospital discharged patients’ medical records. Demographic information, diagnosis, treatment, and discharge status were included in the data. We have used the PEDALFAST dataset which stands for PEDiatric Validation of Variables in Trauma. A survey of patients to find out how they feel about the quality of care they received while they were in the hospital was also a part of the study dataset. The findings of this study will shed light on the ways in which various factors influence the LOS in the hospital and the type of discharge, assisting in the formulation of strategies to enhance the quality and effectiveness of health care delivery.

## Introduction

The aim of our work is to build machine learning models to predict the length of stay (LOS) of a patient in hospital and also their discharge disposition from the hospital based on different patient covariates which are given in this dataset . For this purpose we analyze the PEDALFAST dataset. More details about the dataset as described by the original authors are reproduced below for the reader’s convenience.

A prospective cohort study was carried out at multiple freestanding level I Pediatric Trauma Centers affiliated with the American College of Surgeons as part of the PEDALFAST (PEDiatric Validation of Variables in Trauma) project. Patients under the age of 18 who were admitted to the intensive care unit (ICU) within the first 24 hours with a diagnosis of acute traumatic brain injury (TBI) and a Glasgow Coma Scale (GCS) score of 12 or who had undergone neurosurgical treatment (intracranial pressure [ICP] monitor, external ventricular drain [EVD], craniotomy, or craniectomy) are included in the cohort.

The University of Colorado Denver’s REDCap electronic data capture tools were used to collect and manage the PEDALFAST study data. (Harris and co. 2009) REDCap (Research Electronic Data Capture) is a safe, web-based application that supports the collection of data for research studies. It offers 1) a user-friendly interface for entering validated data; 2) audit trails for monitoring export and data manipulation procedures; 3) automated export procedures that make it easy to download data to common statistical programs; and 4) ways to import data from outside sources.

To build a deep machine learning (ML) model for predicting the LOS and disposition of a patient in the hospital using PEDALFAST data, we first preprocess the data by cleaning and formatting it appropriately to prepare the input to the ML model. Next, we choose a suitable deep learning architecture and train the model on the preprocessed data. Finally, we evaluate the performance of the model using appropriate metrics, such as accuracy and F1 score, and fine-tune it as necessary to improve its performance. We built the models using the Python package H2O. We use H2O.AutoML to prepare multiple models to predict hospital LOS and also hospital disposition. The sections below contain more details about our methods.

## Methodology

In the preprocessing step for the disposition column vector, we converted the original/raw string values to integers and then encoded the values as 1 for mortality/death and 0 for the rest of the disposition outcomes. This approach helps us to understand and predict the mortality rate of the patients at a particular hospital using a binary classification model which predicts ‘death’ or ‘no death’ as two potential outcomes.

We built two models for the LOS prediction. The first model is a classification model that classifies if the patient will have a prolonged stay or a short stay at the hospital for this the short stay is if the LOS is less than mean and prolonged stay is if the LOS is more than the mean (i.e. binary classification for longer or shorter length of stay). The second model is a regression model. In preprocessing of LOS for the classification model, we first found the mean of the LOS and then compared the length of stays of each with the mean and if LOS is more than the mean we give it a score of 1 and if it’s less we give it a score of 0. This way we can find how long a patient is staying in hospital and what is their recovery time. For the regression model, no such preprocessing was required since the model directly estimates the LOS number of days.

We then go through the dataset carefully and drop or remove columns which have NaN values, string values etc. We then perform a test and train split by allocating 80% of the data to the training set and the rest 20% of the data to the test set. After the test-train split, we pass the data to our H2O AutoML model for building multiple models. The H2O machine learning platform’s H2O AutoML feature automates the creation and tuning of machine learning models. It finds the best model for a given dataset by combining methods like grid search, random search, and Bayesian optimization. It is compatible with a wide range of models, including gradient boosting machines, generalized linear models, and deep learning neural networks. Data scientists will be able to quickly identify the best model for their problem without having to manually tune and experiment with various model configurations thanks to this feature, which is intended to save them time and effort. Fig 1 represents a basic architecture of H2O AutoML.

**Fig 1.**
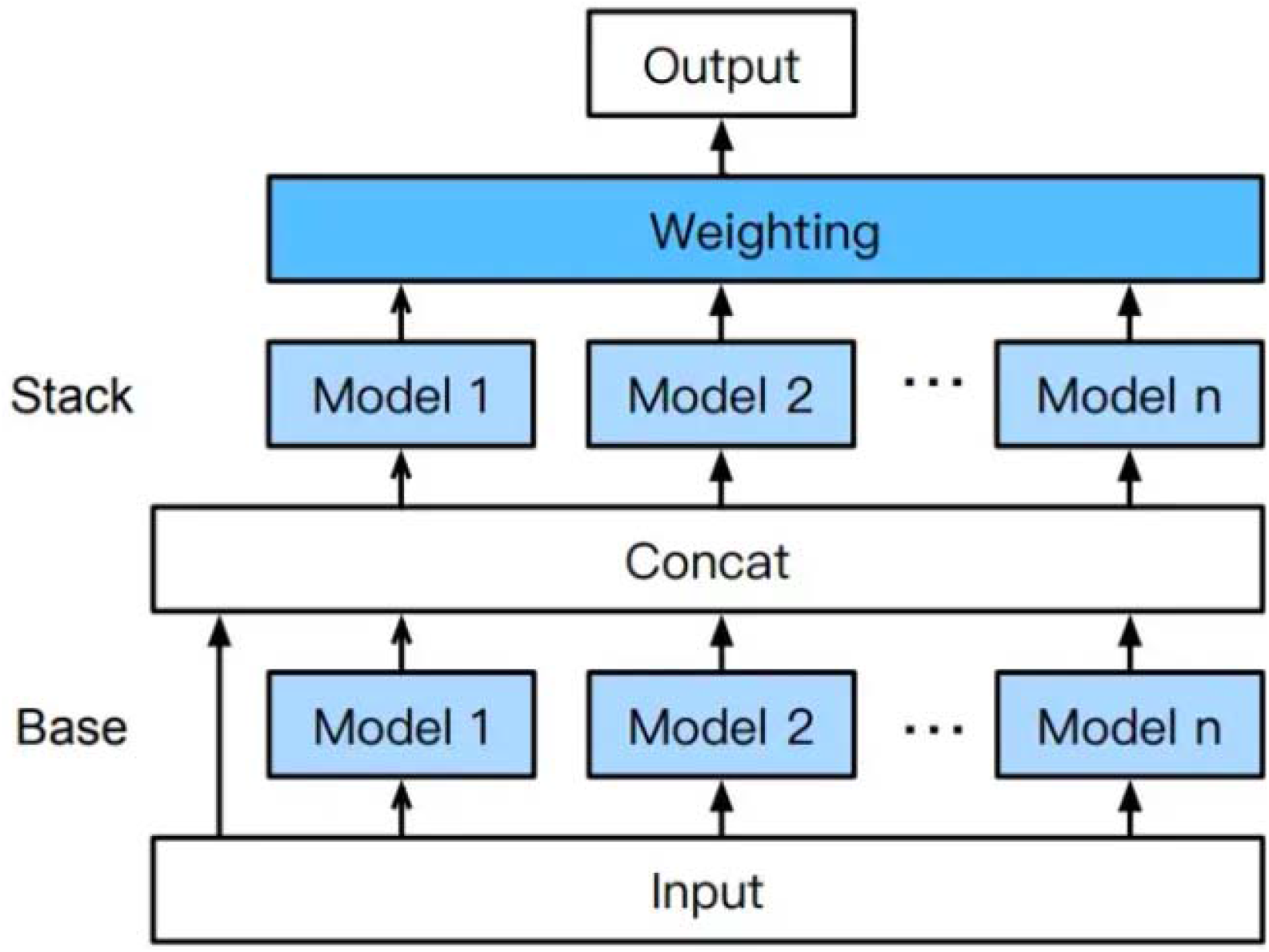
H2O AutoML Architecture

## Exploratory data analysis

In our experiment we are using the PEDALFAST dataset which has a number of columns. We will be discussing some of the important columns used for Hosp.disposition and Hosp.Loss experiments .

### Hosp.disposition EDA

Entnutyn. admittoentnut, gcsicu, hosplos, puplrcticu, cardiacarresticu, cardicarrestyn, admittoicudc1, gcsmotoric, admittocathstart2 etc . also the dataset has the following as output of disposition ‘Inpatient Rehab’, ‘Home, no new supports’, ‘Other’,’Discharge TO or WITH Hospice’, ‘Mortality’,’Short-term Nursing Facility’, ‘Home with Skilled Nursing’ but in our experiment we treat mortality as 1 and the remaining outputs as 0 .

Entnutyn : It means Did the patient receive enteral nutrition or not . If a patient received nutrition is marked as 1 an if the patient did not receive it it is marked as 0 . Out of 388 patients in the data set 333 received the enteral nutrition and 55 didn’t receive enteral nutrition.

Admittoentnut: It Indicates the Days from admission to enteral nutrition . More than 90% of patients it was less than 5 days .

Gcsicu: It is the sum of all gcs’s gcseyeicu, gcsverbalicu and gcsmotoricu . Here GCS is Glasglow coma scale gcseyeicu indicates the reading taken after diagnosing eye, gcsverbalicu indicates the readings taken after having a verbal communication with patient and gcsmotoricu indicates the readings taken after seeing the movement of patient

Hosplos: It tells us about the length of stay of a patient in the hospital or after how many days after admission the patient was discharged or worst case decreased.

Puplrcticu : It tells us about Pupillary reaction on ICU admission . Out of 388 patients there was Both Fixed reaction in 62 patients, Both Reactive reaction in 284 patients and reaction in One Fixed 11 patients.

Cardiacarresitcu : did the patient get Cardiac arrest in ICU or not . Out of all our patients 16 of them suffered cardiac arrest in the ICU.

Cardicarrestyn: Did the patient have a cardiac arrest at any time from the time of injury to the time of hospital discharge.Out of all our patients 59 . patient had a cardiac arrest at any time from the time of injury to the time of hospital discharge

admittoicudc1 : It indicates the Days from admit to ICU discharge for the initial ICU admission.

Gcsmotoricu: indicates the readings taken after seeing the movement of a patient.

Admittocathstart2: Days from admit to second ICU admission (first readmission)

**Table 1.**
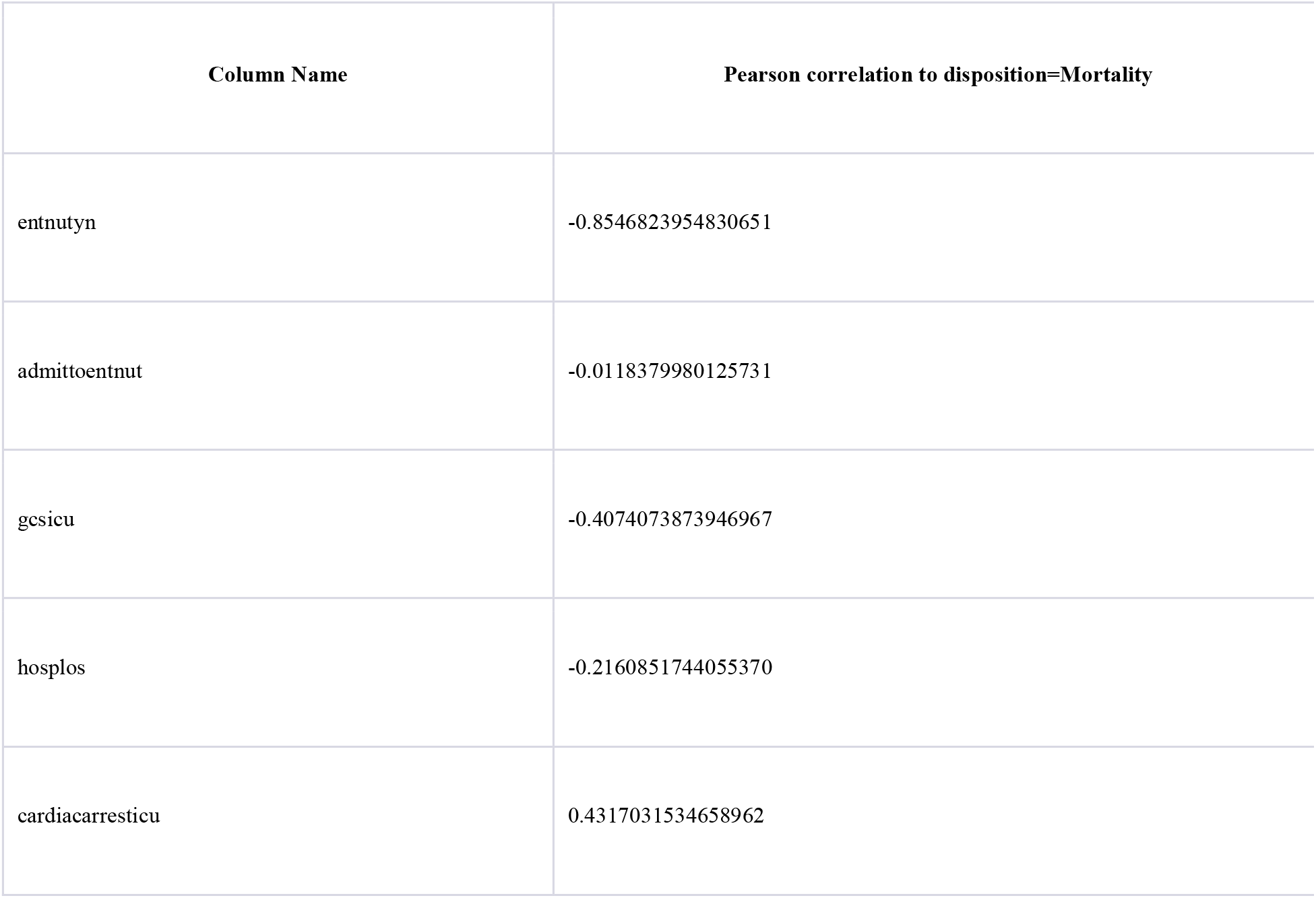
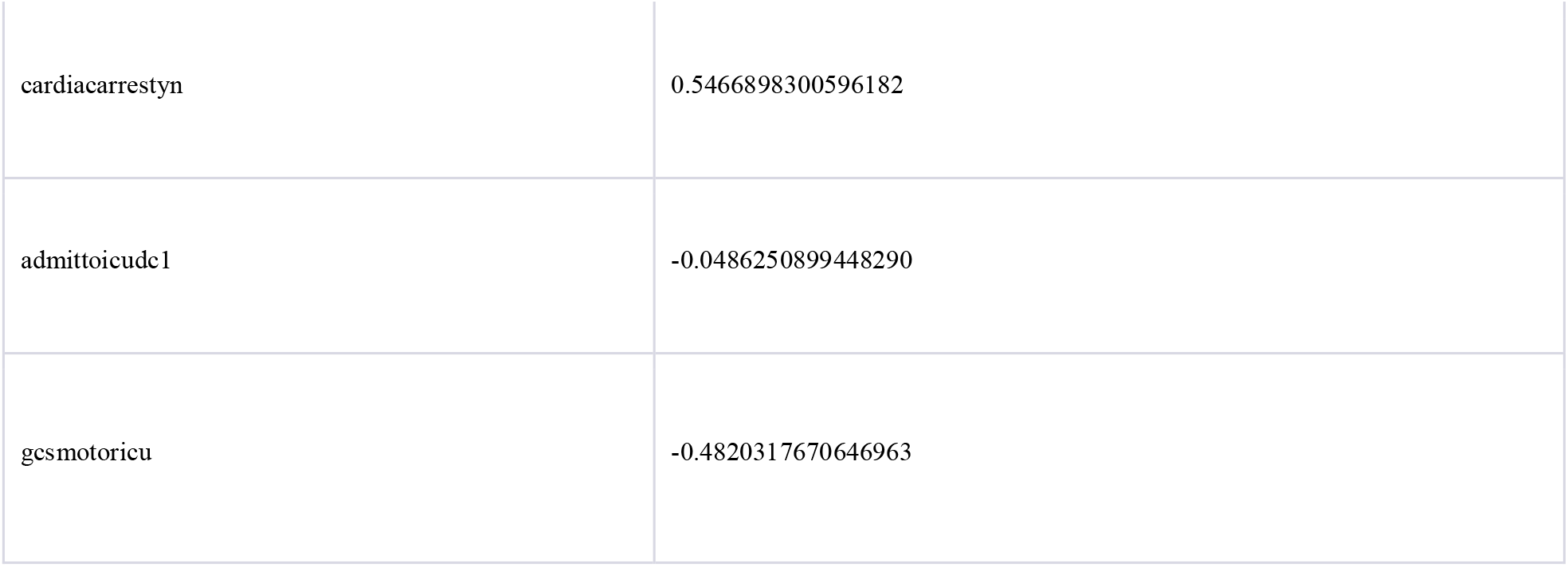
Correlation of variables to Hosp.disposition – From this exploratory data analysis, it can be seen that cardiac events are positively correlated with death, and interventions such as enteral nutrition are negatively correlated with death. Shorter length of stay is correlated with non death.

### Hosp.LOS Exploratory data analysis

newgastyn, icpyn1, cathtype3, icptype1, cathtype2, entnutyn, age, injurymech, sourceinj etc

Newgastyn: it indicates whether the patient received a new gastrostomy or not . Out of 388 patients in the data set 27 received the new gastrostomy and 361 didn’t receive new gastrostomy.

Icpyn1: It Indicates if the patient receives an ICP Monitor or not . Out of 388 patients in the data set 130 received the ICP Monitor and 257 didn’t receive ICP Monitor.

Cathtype3: It tells which Type of third invasive vascular catheter is implanted . Not all the patients had undergone this but Arterial catheter was implanted in 16, Central venous catheter was implanted in 12 patients and Peripherally inserted central catheter (PICC) was implanted in 28.

Icptype1: it tells about the First ICP monitor type . Not all patients were implanted with it but Intraparenchymal (Camino or bolt) was implanted in 92 patients, Ventriculostomy (External Ventricular Drain or EVD) was implanted in 27 and some Other was implanted in 8.

Cathtype2: It tells which Type of Second invasive vascular catheter is implanted . Not all the patients had undergone this but Arterial catheter was implanted in 98, Central venous catheter was implanted in 52 patients and Peripherally inserted central catheter (PICC) was implanted in 16.

Entnutyn : It means Did the patient receive enteral nutrition or not . If a patient received nutrition is marked as 1 and if the patient did not receive it it is marked as 0 . Out of 388 patients in the data set 333 received the nutrition and 55 didn’t receive any nutrition.

Age: It tells us about Age of the patient in days, at time of admission.

Injurymech: It tells us how the patient gets injured . By Fall a total of 72 patients were injured, By Known or suspected abuse around 91 patients were injured, 6 patients were due to Self-harm, 142 due to traffic and 77 due to other miscellaneous reasons .

Sourceinj : It tells us about the Source of Injury Information . The attending Emergency Department (ED) gave information regarding 78 patients, around 136 patients injury information was received from provider note in EMR and Trauma surgery attending surgeons took the information about the rest of 168

**Table 2.** correlation of variables with Hosp.LOS.

| Column Name | Variable correlation with length of stay |
| --- | --- |
| newgastyn | 0.47475897319452265 |
| icpyn1 | 0.40418329160469696 |
| entnutyn | 0.21486666188413014 |
| age | 0.05379095880648816 |

## Results And Analysis

### Hosp.disposition Results

After preprocessing of the data, we trained 50 different models which were based on Gradient Boosting Machine (GBM), Deep Neural Networks (DNN), XGBoost (XGB), Distributed Random Forest (DRF), etc. to predict hosp.disposition. We found that a gradient boosting machine model variant performed the best among the different models. We achieved an area under the curve (AUC) of 0.986424, log.loss of 0.127897, area under precision recall (AUPR) of 0.951531, mean per class error of 0.0889973, root mean square error (RMSE) of 0.180687 and mean square error (MSE) of 0.0326478. All these scores were achieved on the test data which was the 20% of the total data which we split in the preprocessing. Fig 2 represents the confusion matrix for the Hosp.disposition where 0 stands for non - mortality state and 1 stands for mortality, The error is the number of incorrect outputs over the total number of actual outputs .

**Fig 2.**
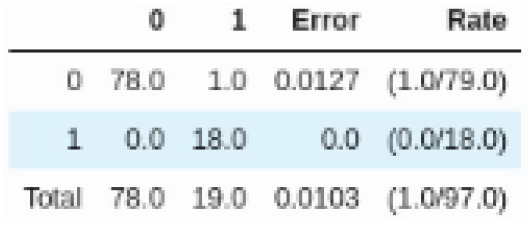
Confusion Matrix for Hosp. disposition

Using the confusion matrix in Fig2 we can calculate many performance metrics which tells us about the performance of our model which are in the table below.

**Table 3.** Performance metrics from Confusion matrix for Hosp.disposition.

| Measure | Value | Derivations |
| --- | --- | --- |
| **Sensitivity** | 1.0000 | $TPR = TP / (TP + FN)$ |
| **Specificity** | 0.9873 | $SPC = TN / (FP + TN)$ |
| **Precision** | 0.9474 | $PPV = TP / (TP + FP)$ |
| **Negative Predictive Value** | 1.0000 | $NPV = TN / (TN + FN)$ |
| **False Positive Rate** | 0.0127 | $FPR = FP / (FP + TN)$ |
| **False Discovery Rate** | 0.0526 | $FDR = FP / (FP + TP)$ |
| **False Negative Rate** | 0.0000 | $FNR = FN / (FN + TP)$ |
| **Accuracy** | 0.9897 | $ACC = (TP + TN) / (P + N)$ |
| **F1 Score** | 0.9730 | $F1 = 2TP / (2TP + FP + FN)$ |

The accuracy of our model is 0.9897. We obtained a recall or sensitivity score of 1.00 which is a measure of how well a classification model correctly identifies positive instances in our case its mortality of the patient . We also got a specificity score of 0.9873 which is a measure of how well a classification model correctly identifies negative instances. The precision score was 0.9474. Overall, the metrics are good on the test dataset. It remains to be seen how well the model generalizes to patient data from other hospitals.

### Variable Importance and Shapley plots for Hosp.disposition

The most significant variables are listed in descending order of mean Gini decrease in the variable importance plot. The top variables contribute more to the model than the bottom variables. We found variables which affect the disposition of patients with the help of the plot given in Fig.3. For variable importance, we used a threshold of 0.4 (i.e. if the variable receives a score of 0.4 or above for Gini decrease then it is considered as important). Let’s discuss in detail what these variables indicate in our dataset . Two variables have crossed the threshold of 0.4 and become the most important variable for Hosp.disposition . The most important variable *entnutyn* tells us whether the patient receives any enteral nutrition or not . This tells that if a patient receives enteral nutrition, the probability of a mortality event decreases. The next variable is *admittoentnut* which tells us about the number of days in the hospital the patient is on enteral nutrition.

**Fig 3.**
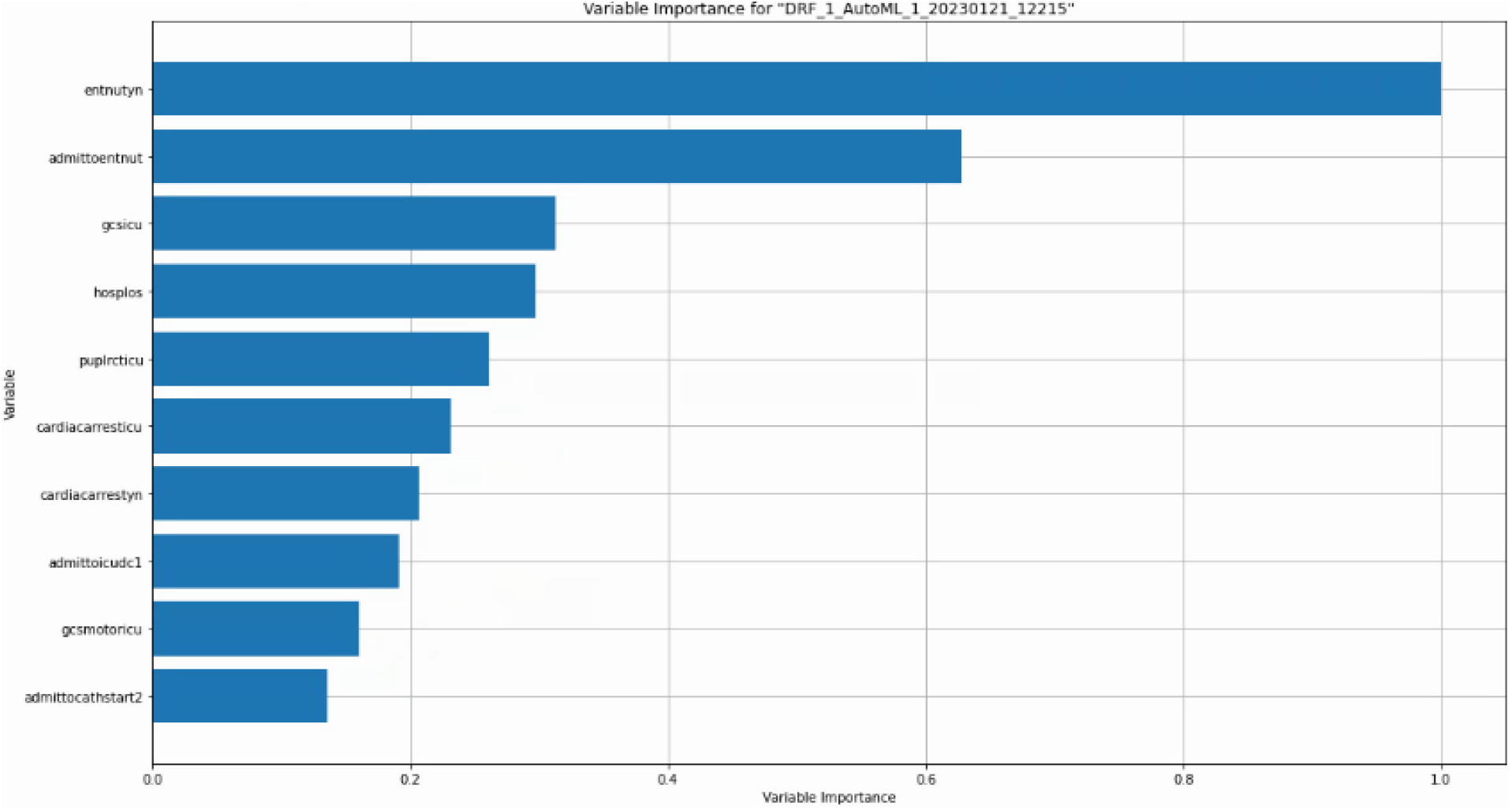
Variable Importance Plot

The summary plot combines feature effects and importance. Shapley values for each instance of each feature can be seen at each point on the summary plot. The feature and each instance’s Shapley value determine the position on the y-axis and x-axis, respectively. We can see that the most important feature, LSTAT, has a wide range of Shapley values. From low to high, the color indicates the feature’s value. We are able to get a sense of the distribution of the Shapley values per feature because overlapping points are jittered in the direction of the y-axis. The features are sorted by how important they are. The Shap Summary plot for Hosp.disposition is shown in Fig.4. From the plot, we can see that the most important feature is again *entnutyn* which represents whether the patient receives any enteral nutrition or not .

**Fig 4.**
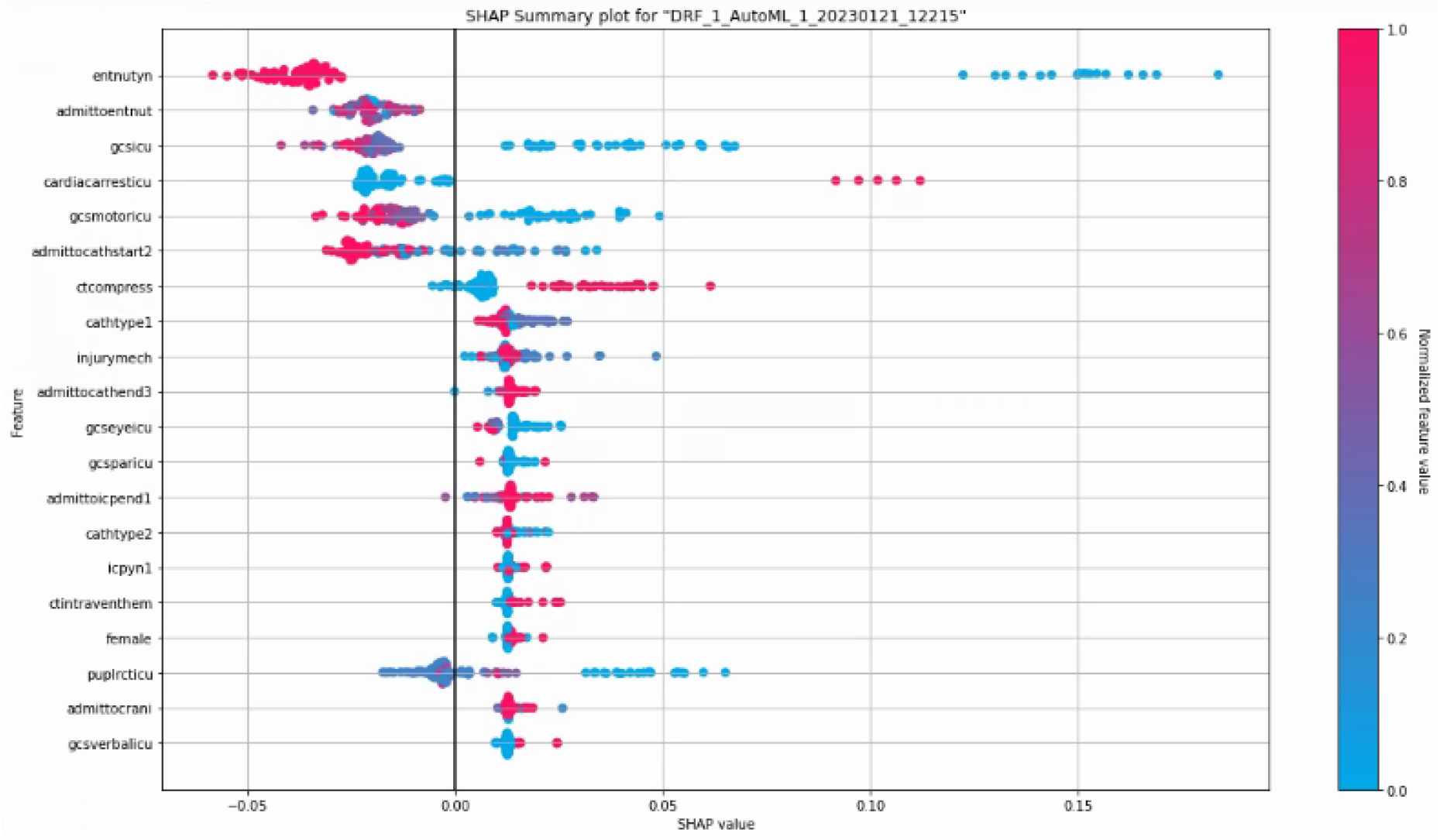
Shap Summary Plot for Hosp.disposition

**Fig 5.**
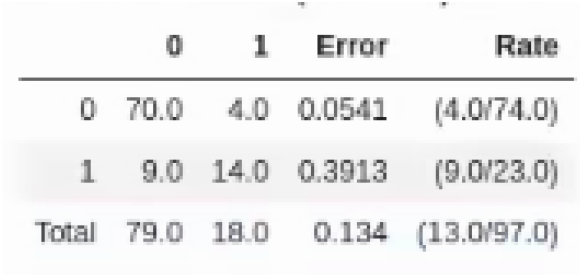
Confusion Matrix for HospLOS

### Hospital LOS classification Results

After finishing the preprocessing of the data we trained around 50 different models which were based on Gradient Boosting Machine (GBM), Deep Neural Networks (DNN), XGBoost (XGB), StackedEnsemble etc, for predicting hospital length of stay. We found that a Gradient Boosting Machine model performed the best among the different models. We achieved an Area under the curve (AUC) of 0.890495, Log.loss of 0.381149, area under precision recall (AUPR) of 0.799785 mean per class error of 0.150934, Root mean square error (RMSE) of 0.34483 and mean square error (MSE) of 0.118908. All these scores were achieved on the test data which was the 20% of the total data which we split in the preprocessing . Fig 3 represents the confusio matrix for the HospLOS.

Using the confusion matrix in Fig.3 we can calculate many performance metrics which tells us about the performance of our model which are in Table below.

**Table 4.** Performance metrics from Confusion matrix for Hosp.LOS.

| Measure | Value | Derivations |
| --- | --- | --- |
| **Sensitivity** | 0.6087 | $TPR = TP / (TP + FN)$ |
| **Specificity** | 0.9459 | $SPC = TN / (FP + TN)$ |
| **Precision** | 0.7778 | $PPV = TP / (TP + FP)$ |
| **Negative Predictive Value** | 0.8861 | $NPV = TN / (TN + FN)$ |
| **False Positive Rate** | 0.0541 | $FPR = FP / (FP + TN)$ |
| **False Discovery Rate** | 0.2222 | $FDR = FP / (FP + TP)$ |
| **False Negative Rate** | 0.3913 | $FNR = FN / (FN + TP)$ |
| **Accuracy** | 0.8660 | $ACC = (TP + TN) / (P + N)$ |
| **F1 Score** | 0.6829 | $F1 = 2TP / (2TP + FP + FN)$ |

The accuracy of our model looking at the confusion matrix is 0.8660. We obtained a recall or sensitivity score of 0.6087 which is a measure of how well a classification model correctly identifies positive instances. We also got a specificity score of 0.9459 and a precision of 0.7778.

### Variable Importance and Shapley plots for Hosp.LOS

The most significant variables are listed in descending order of mean Gini decrease in the variable importance plot. In addition to having superior predictive power, the top variables contribute more to the model than the bottom variables. We also found which variable affects the LOS of patients with the help of the plot given in Fig.6. For the importance, we chose a threshold of at least 0.4, that is if the variable receives a score of 0.4 or above then it is considered as important . The most important variable with the highest importance score is *newgastyn* which indicates whether the patient received any new gastrostomy or not. For nutritional support, a gastrostomy is a procedure in which a gastrostomy tube is inserted into the stomach. This is quite important as in the case of any surgery or illness, if a patient is not in a state to chew their own food (due to which he cant intake the required nutrition). This intervention could also be a proxy indicator of the patient’s state of recovery (i.e. a patient who doesn’t need this intervention is likely recovering faster).

**Fig 6.**
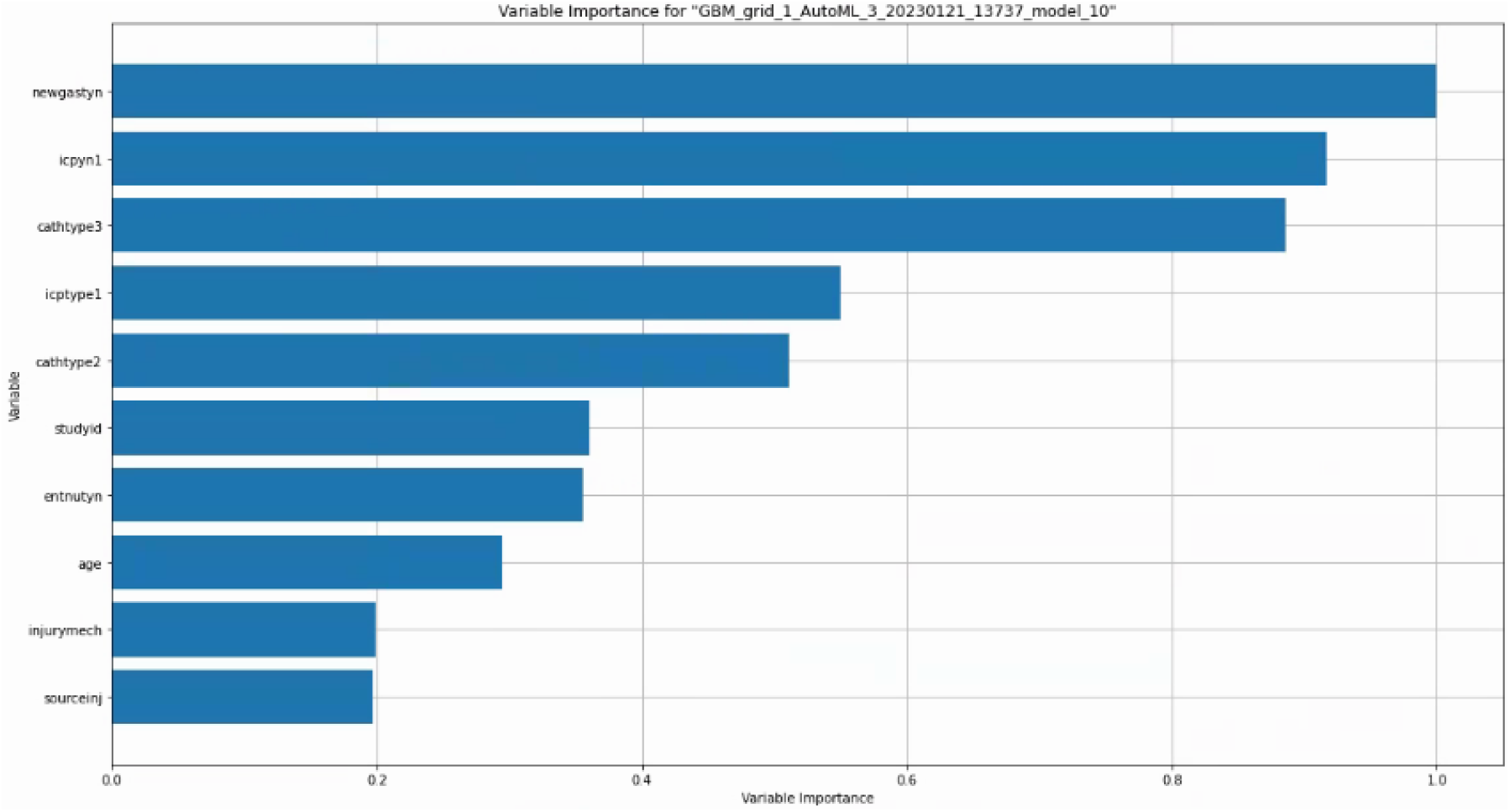
Variable Importance Plot

The next variable is *icpyn1* which tells us if the patient was monitored using an ICP monitor which is used to monitor intracranial pressure. This helps the doctors figure out if the symptoms are caused by high or low pressure in the cerebrospinal fluid (CSF). A small, pressure-sensitive probe that is inserted through the skull is used to directly measure the pressure in the patient’s head during the test. This is crucial information for diagnosis purposes as it helps the doctor in making decisions .

The summary plot combines feature effects and importance. Shapley values for each instance of each feature can be seen at each point on the summary plot. The feature and each instance’s Shapley value determine the position on the y-axis and x-axis, respectively. You can see that the most important feature, LSTAT, has a wide range of Shapley values. From low to high, the color indicates the feature’s value. We are able to get a sense of the distribution of the Shapley values per feature because overlapping points are jittered in the direction of the y-axis. The features are sorted by how important they are. The Shap Summary plot for HospLOS is shown in Fig.7. From the plot below, we can see that for *entnutyn, cathtype3, icpyn1* and *newgastyn* are well separated in blue versus red color comparison of shap summary plot . *entnutyn* represents whether the patient receives any enteral nutrition or not. *cathtype3* tells us about the type of third invasive vascular catheters. *icpyn1* tells us if the patient was monitored using an ICP monitor which is used to monitor intracranial pressure. *newgastyn* indicates whether the patient received any new gastrostomy or not

**Fig 7.**
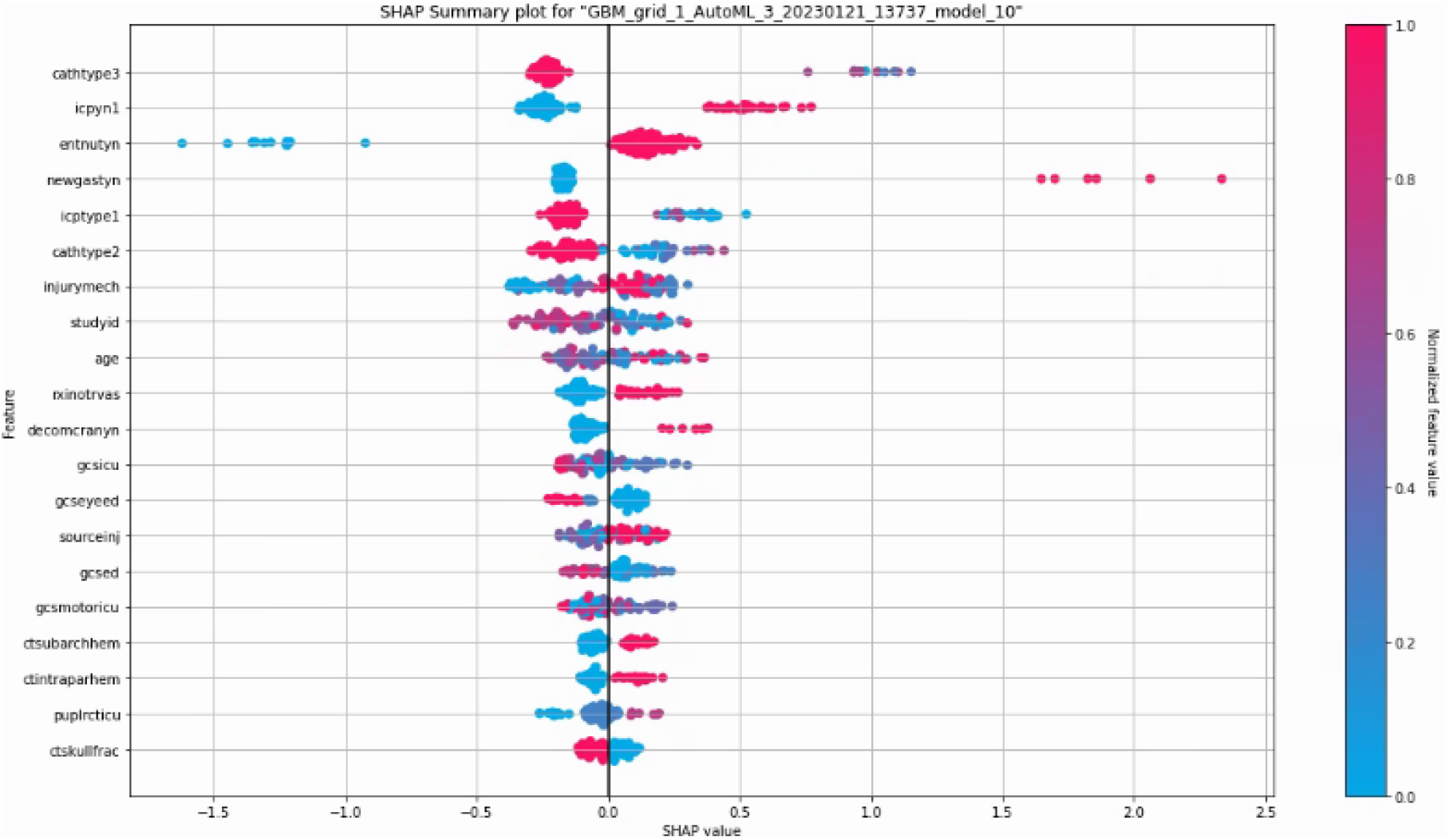
Shap Summary Plot For HospLOS

### Hosp.LOS Regression Results

After finishing the preprocessing of the data we trained around 50 different models which were based on Stack Ensemble, Gradient Boosting Machine (GBM), Deep Neural Networks (DNN) and XGBoost (XGB), we found that a Gradient Boosting Machine model performed the best among the different models for this particular data . We achieved mean residual deviance of 482.06, Root mean squared error (RMSE) of 21.9559, Mean squared error (MSE) of 482.06, Mean Absolute Error (MAE) of 10.5301 which means that there is a 10 days error for LOS .All these scores were achieved on the test data which was the 20% of the total data which we split in the preprocessing

## Conclusion

A novel model was developed in this paper to predict hospital disposition and hospital length of stay (LOS) based on a variety of patient and intervention related parameters. We use H2O AutoML for this, which enables us to train multiple models simultaneously and returns the best one. The performance metric for each model is then generated. Based on the findings and analysis, we can conclude that we have developed a robust deep learning model for predicting both the LOS and the discharge type for hospital patients. However, it remains to be seen how well this model may generalize, and how much it will need to be fine tuned for good performance on data from other hospitals from different locations.

## Data Availability

All data is available publicly in the PEDALFAST R package

